# Targeted screening for Alzheimer’s disease clinical trials using data-driven disease progression models

**DOI:** 10.1101/2021.01.29.21250773

**Authors:** Neil P. Oxtoby, Cameron Shand, David M. Cash, Daniel C. Alexander, Frederik Barkhof, for the Alzheimer’s Disease Neuroimaging Initiative, the Alzheimer’s Disease Cooperative Study

## Abstract

Heterogeneity in Alzheimer’s disease progression contributes to the ongoing failure to demonstrate efficacy of putative disease-modifying therapeutics that have been trialled over the past two decades. Any treatment effect present in a subgroup of trial participants (responders) can be diluted by non-responders who ideally should have been screened out of the trial. How to identify (screen-in) the most likely potential responders is an important question that is still without an answer. Here we pilot a computational screening tool that leverages recent advances in data-driven disease progression modelling to improve stratification. This aims to increase the sensitivity to treatment effect by screening out non-responders, which will ultimately reduce the size, duration, and cost of a clinical trial. We demonstrate the concept of such a computational screening tool by retrospectively analysing a completed double-blind clinical trial of donepezil in people with amnestic mild cognitive impairment (clinicaltrials.gov: NCT00000173), identifying a data-driven subgroup having more severe cognitive impairment who showed clearer treatment response than observed for the full cohort.

## 1 INTRODUCTION

Alzheimer’s Disease (AD) is one of the most important socioeconomic challenges of the 21st century, being the leading cause of age-related dementia in an ageing global population. Despite decades of research and clinical trials of potential therapies (1), no trials have been able to prove disease-modifying efficacy (2, 3, 4, 5, 6, 7). There are multiple possible explanations for this. For example, potentially targeting the “wrong” pathology at the wrong time — typically amyloid protein pathogens are the target but if a treatment is given to symptomatic individuals, it may be too late to halt or reverse any damage done. Notwithstanding this, enrolling the right people at the right time (disease stage) into a clinical trial remains a considerable challenge because of undetected heterogeneity in phenotype/presentation (8) and/or ensuring the underlying pathology is present (9), which can be a general problem because clinical trials often cannot adapt their designs to accommodate research discoveries made after they have begun. This can result in enrolment of non-responders into a clinical trial that wash out treatment effect in any subgroup of responders. Identification of non-responders typically occurs in post hoc subgroup analysis, which does not confer the benefits of a reduced trial size, and requires careful analysis to infer conclusions which can be misleading (10, 11). Given the breadth of evidence in support of the amyloid hypothesis (12) that has driven this clinical research for two decades, albeit with some controversies (13), here we focus on the aforementioned challenges of screening to identify the right participants at the right time. The good news is that there has been a swell of computational research into unravelling the heterogeneity of Alzheimer’s disease progression over the past decade (e.g., see (14)), driven largely by the increasing availability of large open medical datasets (15).

Computational approaches for ageing and age-related diseases have been designed to fuse multimodal data into a quantitative template (16) of disease progression. These signatures often include a patient staging mechanism (17) that provides a quantitative tool for fine-grained, individualised inference based on disease severity that goes above and beyond standard clinical phenotyping using patient symptoms. A recent innovation of data-driven disease progression modelling incorporates unsupervised machine learning, i.e., clustering, to provide both subtype and stage inference (18). A frequent occurrence in this literature are claims of how these data-driven models can benefit clinical trials in Alzheimer’s disease, but we are yet to find any evidence of studies actually analysing clinical trial data to demonstrate the claimed benefit.

In this work we demonstrate the potential of data-driven models of disease progression to enhance clinical trials in Alzheimer’s disease via targeted screening. We achieve this by example, using a particular modelling approach — the event-based model (19) — in a *post hoc* subgroup analysis of a particular completed clinical trial that concluded without evidence of efficacy (20).

## 2 MATERIALS AND METHODS

This section describes the data, the computational model, and the statistical analysis used in our study. Overall, our analysis includes three steps. First, we fit a data-driven disease progression model of cognitive decline in AD to data from a large multicentre observational study, the Alzheimer’s Disease Neuroimaging Initiative (ADNI; *training set*). Second, we use this computational model to score disease progression at baseline for participants in the completed “MCI” clinical trial from the Alzheimer’s Disease Cooperative Study (ADCS-MCI; *test set*). Finally, this disease progression score is used to stratify the ADCS-MCI Trial participants for a *post hoc* analysis of subgroup treatment effect.

### 2.1 Data

Our reference model fit to data from the ADNI observational study is used to stage participants from the ADCS-MCI clinical trial (clinicaltrials.gov: NCT00000173; (20)). For this we use a set of features common to both data sets, which is a subset of cognitive instruments used in the ADCS-MCI trial (see the vertical axis of Results Figure 1), taking care to exclude ADAS-Cog (being a secondary outcome of the trial).^1^ For simplicity, we included only ADNI participants having complete data for this feature set.

**Figure 1.**
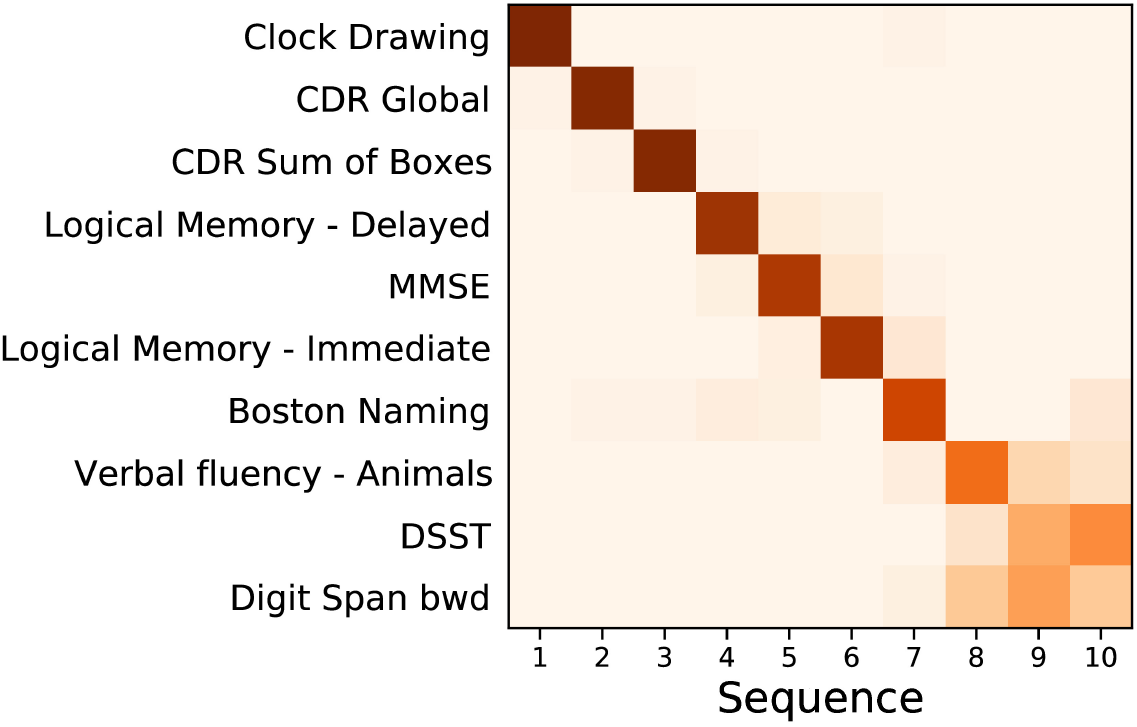
Event based model of cognitive decline (ADNI). Positional density/variance diagram showing the sequence (top to bottom) and uncertainty (left to right) under 5-fold cross-validation (repeated 10 times). Abbreviations: CDR — clinical dementia rating; MMSE — mini-mental state examination; bwd — backward; DSST — digit symbol substitution test.

Data used in the preparation of this article were obtained from the Alzheimer’s Disease Neuroimaging Initiative (ADNI) database (adni.loni.usc.edu). The ADNI was launched in 2003 as a public-private partnership, led by Principal Investigator Michael W. Weiner, MD. The primary goal of ADNI has been to test whether serial magnetic resonance imaging (MRI), positron emission tomography (PET), other biological markers, and clinical and neuropsychological assessment can be combined to measure the progression of mild cognitive impairment (MCI) and early Alzheimer’s disease (AD).

Additional data used in the preparation of this article were obtained from the Alzheimer’s Disease Cooperative Study (ADCS) database (adcs.org). Specifically, we analyse data from the completed ADCS-MCI clinical trial of donepezil and vitamin E, reported in Petersen et al. (20). The ADCS-MCI trial aimed to assess the efficacy of vitamin E and donepezil in subjects with amnestic MCI. The primary end point was the time to the development of possible or probable AD dementia, with secondary outcomes on cognition and function. Measurements were taken at 6-month intervals until the end of the trial (36 months). At screening, 769 subjects were included in the trial, randomized into 259, 257, and 253 subjects for the placebo, vitamin E, and donepezil arms, respectively — reducing to 174, 158, and 145 by the end of the trial.

### 2.2 Event-based Model

The event-based model (EBM) (17, 19) estimates the most likely sequence, and uncertainty in this sequence, of observable events in the pathophysiological cascade (21) of a progressive disease. This is achieved directly from the data distributions in diseased and healthy groups and without *a priori*-defined disease stages or biomarker cutpoints. The EBM, in its various versions, has been applied to a variety of diseases since 2011, e.g., (19, 8, 22, 23, 24, 25). For a detailed intuitive description of the EBM, we refer the reader to (22).

Here we employ the recently-developed kernel density estimation (KDE) EBM that copes naturally with the ceiling/floor effects seen in cognitive data (8), and gives a cleaner interpretation of the model by exploiting prior information on disease direction (22). To improve generalizability, we perform repeated 5-fold cross-validation (10 repeats) and combine all 50 sets of posterior samples of the EBM into a cross-validated positional density map (22).

The EBM affords us a screening tool by way of the patient staging mechanism introduced by Young, *et al.* (17). This process assigns a model stage (disease progression score) that maximizes the likelihood given an individual’s set of measurements. Here we use the ADNI-trained EBM to stage baseline data from the ADCS-MCI clinical trial, then stratify subjects into strata based on disease progression scores for *post hoc* subgroup analyses. In future, this process could be performed as part of the screening process to homogenize the clinical trial cohort.

### 2.3 Statistical Analysis

Our hypothesis is that AD clinical trial cohorts are likely to contain undetected heterogeneity that washes out treatment effects which may exist in an independently identifiable subgroup of responders. Accordingly, in order to examine whether our proposed screening tool can detect this heterogeneity and reveal such a subgroup of responders, our *post hoc* subgroup analysis of the ADCS-MCI clinical trial closely follows the primary analyses in (20). We describe the key steps below.

#### Primary outcome

We use Kaplan-Meier estimators to estimate the rate of progression from MCI to AD over the course of the trial. Additionally, Cox proportional-hazards models were constructed to compare the risk for progression in each treatment arm with the placebo (using baseline age, MMSE, and APOE-*E*4 carrier status as covariates). This intention-to-treat analysis in the trial was conducted for both placebo vs. vitamin E and placebo vs. donepezil, but in this paper we focus on the latter.

To correct for multiple comparisons in the Cox proportional-hazards model (for the two treatment arms), the Hochberg method was used. As our introduction of subgroups increases the number of comparisons made, we extend this adjustment for the total number of subgroups, regardless of whether a single subgroup is the focus of analysis.

#### Secondary outcome

We compare ADAS-Cog 13 scores between placebo and donepezil arms in subgroups at each 6-month interval to assess the difference in longitudinal cognitive decline. A two-sided Mann-Whitney U-test is used to compare the treatment groups at each time point for each subgroup, correcting for multiple comparisons using the Hochberg method.

## 3 RESULTS

### 3.1 Reference Model

Figure 1 shows a positional variance diagram for an event-based model (8) of cognitive decline due to probable Alzheimer’s disease, across a set of cognitive instruments from *N* = 810 (of 2040) ADNI participants (229 cognitively normal (CN), 181 AD, 400 MCI) having complete data (see Methods). The cross-validated model’s confidence in the sequence is higher where the positional variance is reduced — a dark diagonal corresponds to strong confidence in the data-driven ordering. The estimated sequence of cognitive decline starts from the Clock Drawing test and Clinical Dementia Rating (CDR), through tests of memory recall (Logical Memory) and general cognition (MMSE), to verbal fluency (Boston Naming; Animals), working memory (Digit span backwards), and executive function (Digit Symbol Substitution Test, DSST).

Figure 2 shows a key component of the EBM — the normal/abnormal mixture models for each cognitive instrument (blue/orange solid lines, respectively), and the resulting cumulative probability of an event having occurred (dashed lines) (19). These sigmoidal event probabilities quantify divergence from normality (22) and provide a visualisation of the data-driven event threshold (akin to a data-driven biomarker cutpoint). Histograms show the AD (orange) and CN (blue) data from ADNI. Early/late events are, respectively, those that have occurred in many/few patients and thus show greater/smaller separation between the group histograms.

**Figure 2.**
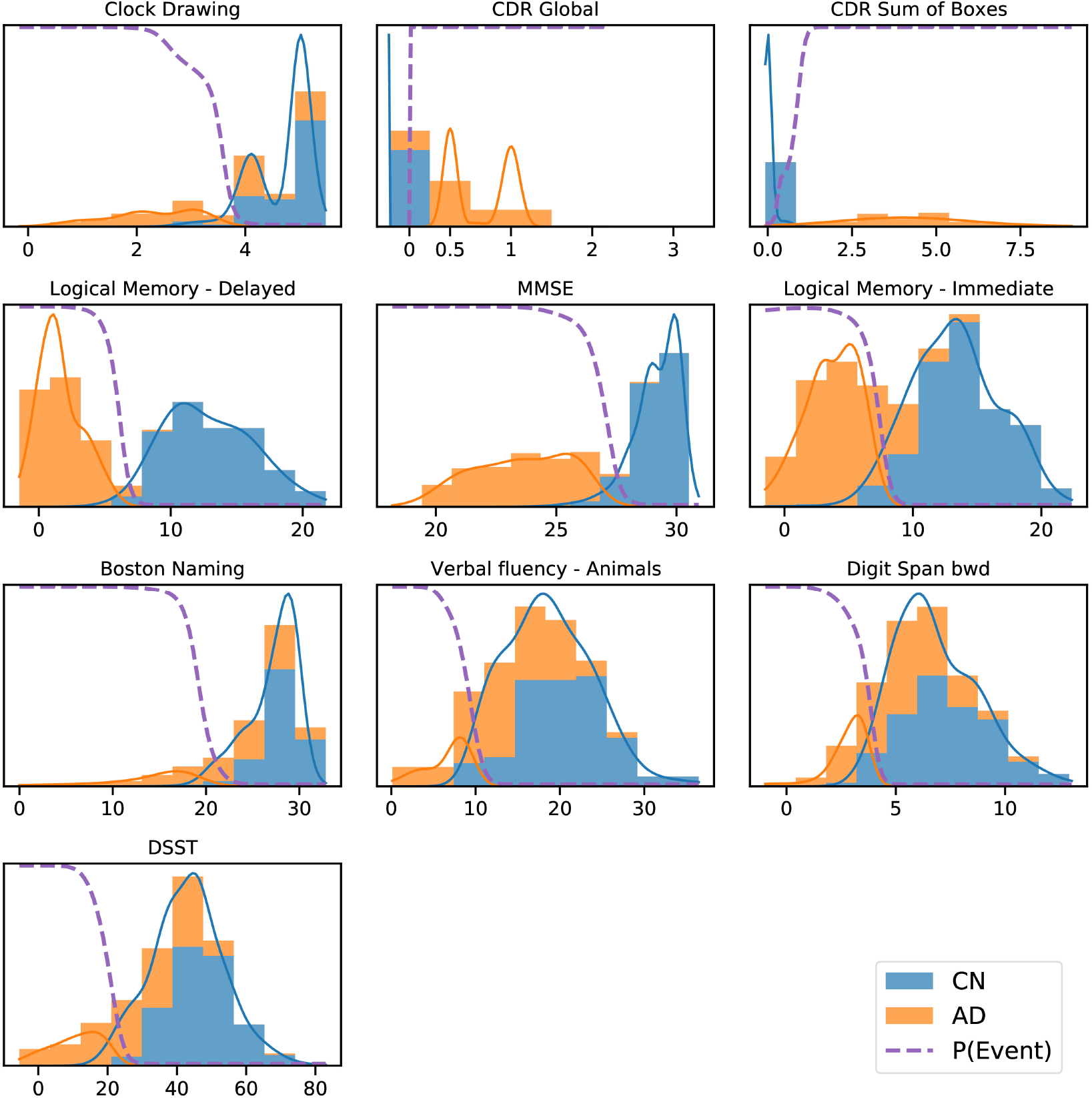
ADNI data histograms (adjusted for age and education level) and EBM mixture models for each feature. Orange bars corresponds to AD patient data, blue bars to data from CN participants, showing the “normal” and “abnormal” distributions and the determined probability of the event having occurred (dashed line).

### 3.2 Patient staging: re-screening the ADCS-MCI trial

Figure 3 shows the distribution of patient stages assigned to participants in the (**A**) ADNI study and (**B**) ADCS-MCI trial, using the ADNI-trained EBM shown in Figure 1. The MCI distributions show considerable heterogeneity, with a notable late-stage ADCS-MCI subgroup beyond stage 8 in Figure 3**B**, delineated by a red dashed line. Table 1 compares the whole ADCS-MCI cohort and 2 subgroups (“Late-stage” and “Others”) on demographic and cognitive measures at baseline.

**Table 1.** Demographic and Cognitive comparison of ADCS-MCI trial participants (All) and the model-determined subgroups thereof (“Late-stage” and “Others”).

| Measure | Group |  |  |
| --- | --- | --- | --- |
|  | All<br>(N=769) | Others<br>(N=648) | Late-stage<br>(N=121) |
| Age (years) | 72.9 (7.3) | 73.0 (7.2) | 72.4 (7.9) |
| Education (years) | 14.6 (3.1) | 14.6 (3.1) | 15.0 (3.0) |
| Sex (% female) | 352 (45.8%) | 290 (44.8%) | 62 (51.2%) |
| APOE-ε4 carrier (%) | 424 (55.1%) | 352 (54.3%) | 72 (59.5%) |
| Donepezil Arm (%) | 253 (32.9%) | 219 (33.8%) | 34 (28.1%) |
| Vitamin E Arm (%) | 257 (33.4%) | 216 (33.3%) | 41 (33.9%) |
| Placebo Arm (%) | 259 (33.7%) | 213 (32.9%) | 46 (38.0%) |
| ADAS-Cog 11 | 11.3 (4.4) | 10.8 (4.2) | 14.1 (4.0) |
| ADAS-Cog 13 | 17.7 (6.1) | 17.0 (5.9) | 21.6 (5.6) |
| ADAS-Cog Q4 | 6.3 (2.2) | 6.1 (2.2) | 7.3 (2.0) |
| Boston Naming | 6.9 (2.4) | 7.3 (2.2) | 5.1 (2.5) |
| CDR Global | 0.5 (0.0) | 0.5 (0.0) | 0.5 (0.0) |
| CDR Sum of Boxes | 1.8 (0.8) | 1.8 (0.8) | 2.2 (0.8) |
| Clock Drawing | 4.3 (0.9) | 4.5 (0.8) | 3.4 (1.0) |
| Digit Span bwd | 6.2 (2.1) | 6.4 (2.1) | 5.1 (1.9) |
| DSST | 31.5 (10.9) | 33.4 (10.2) | 21.1 (8.0) |
| Logical Memory - Delayed | 3.3 (2.4) | 3.5 (2.5) | 2.2 (2.0) |
| Logical Memory - Immediate | 6.2 (3.1) | 6.5 (3.1) | 4.7 (2.7) |
| MMSE | 27.3 (1.8) | 27.5 (1.8) | 26.2 (1.7) |
| Verbal fluency - Animals | 15.8 (5.2) | 16.8 (5.0) | 10.5 (3.0) |

**Figure 3.**
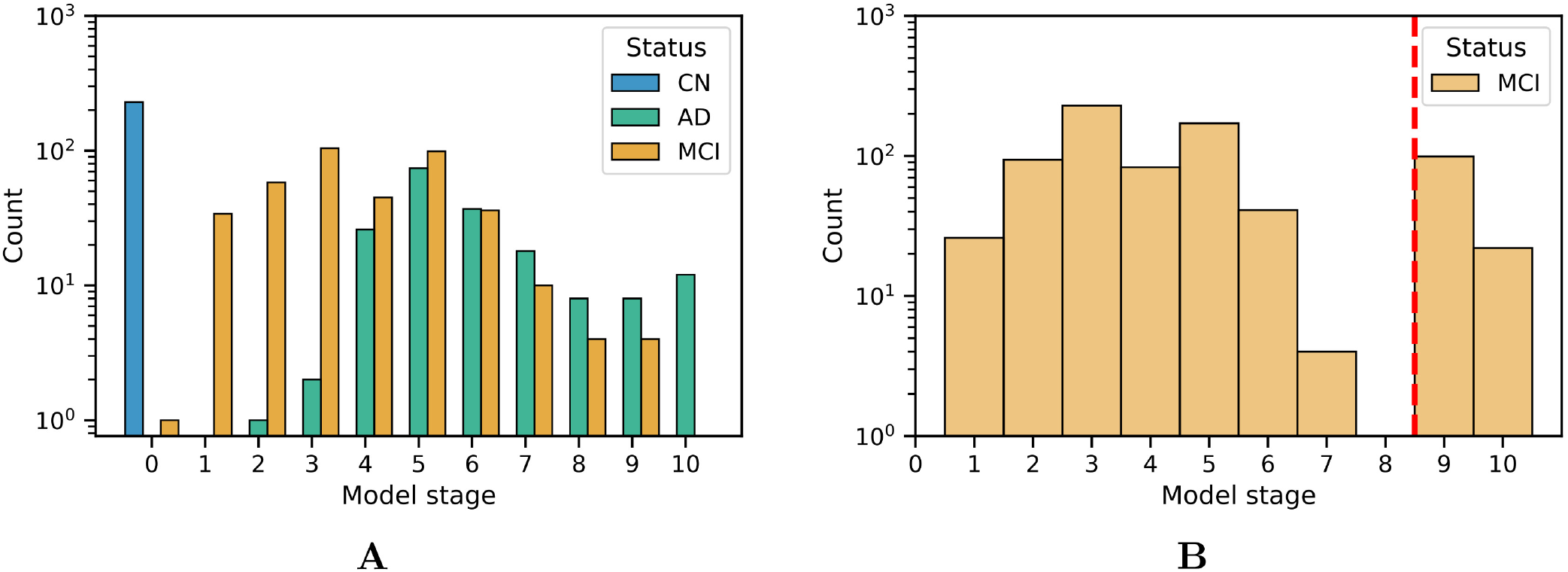
Histograms of model stage for subjects in the ADNI dataset (**A**) and ADCS-MCI trial (**B**).

#### Primary outcome

Figure 4 shows Kaplan-Meier curves for the whole ADCS-MCI cohort (**A**), the early-to-middle “Others” subgroup (**B**) and the “Late-stage” subgroup (**C**) in the placebo and donepezil arms, illustrating the change in survival rates (specifically, not progressing to probable AD dementia) during the trial. For each survival function estimate, 95% confidence intervals are shown in the shaded area. Figure 5 shows the corresponding hazard ratios and 95% confidence intervals for Cox proportional-hazards models quantifying the risk of progression from MCI to AD. Although there are no significant differences between all subjects (hazard ratio 0.80; 95% CI 0.57–1.13; p=0.42), the estimated effect seems larger than in the early-to-middle stage subgroup (hazard ratio 1.00; 95% CI 0.67–1.51; p=0.99), or the late-stage subgroup (hazard ratio 0.55; 95% CI 0.28–1.07; p=0.24).

**Figure 4.**
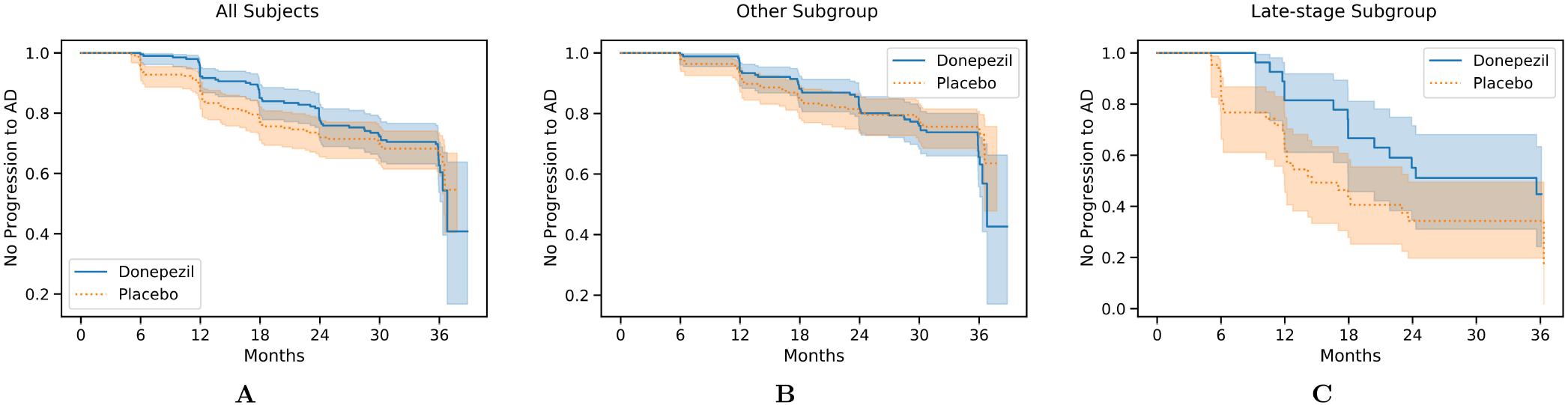
Kaplan-Meier survival curves for all 769 participants (**A**), the “Others” subgroup (**B**), and the “Late-stage” subgroup (**C**) in the ADCS-MCI trial.

**Figure 5.**
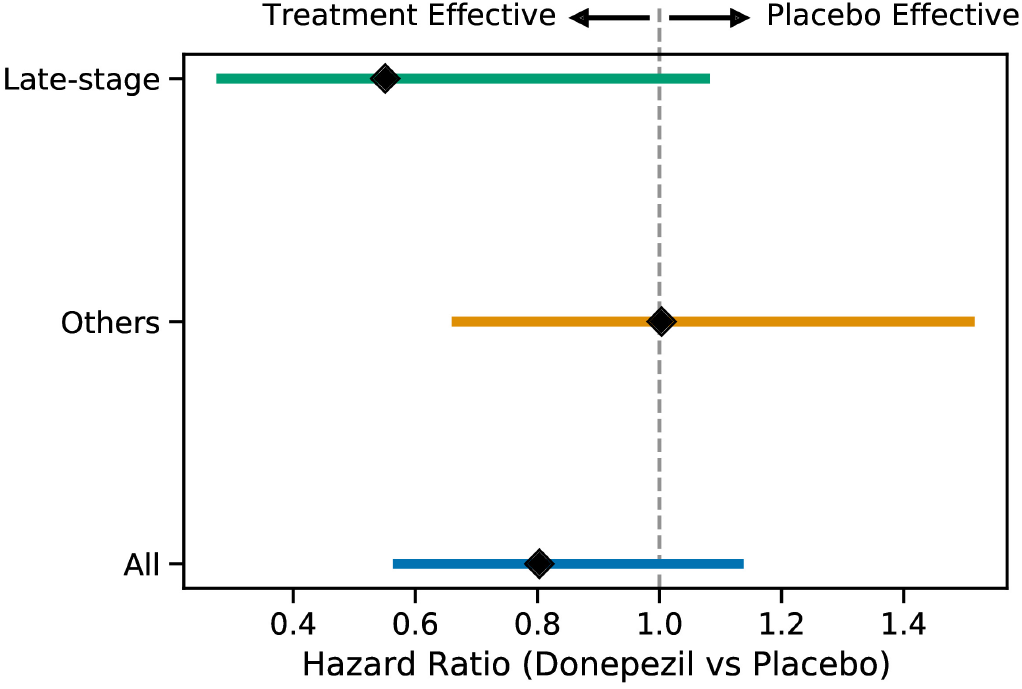
Hazard ratios (with 95% confidence intervals) for the progression to AD for the two subgroups and all subjects when comparing the placebo and donepezil arms.

Figure 6 shows ADAS-Cog 13 scores at 6-month intervals throughout the ADCS-MCI trial separately for the two subgroups. Conducting a two-sided Mann-Whitney U-test at each time point, no significant difference (in adjusted p-values) was found in either subgroup, despite the apparent trend towards treatment effect in the late-stage subgroup.

**Figure 6.**
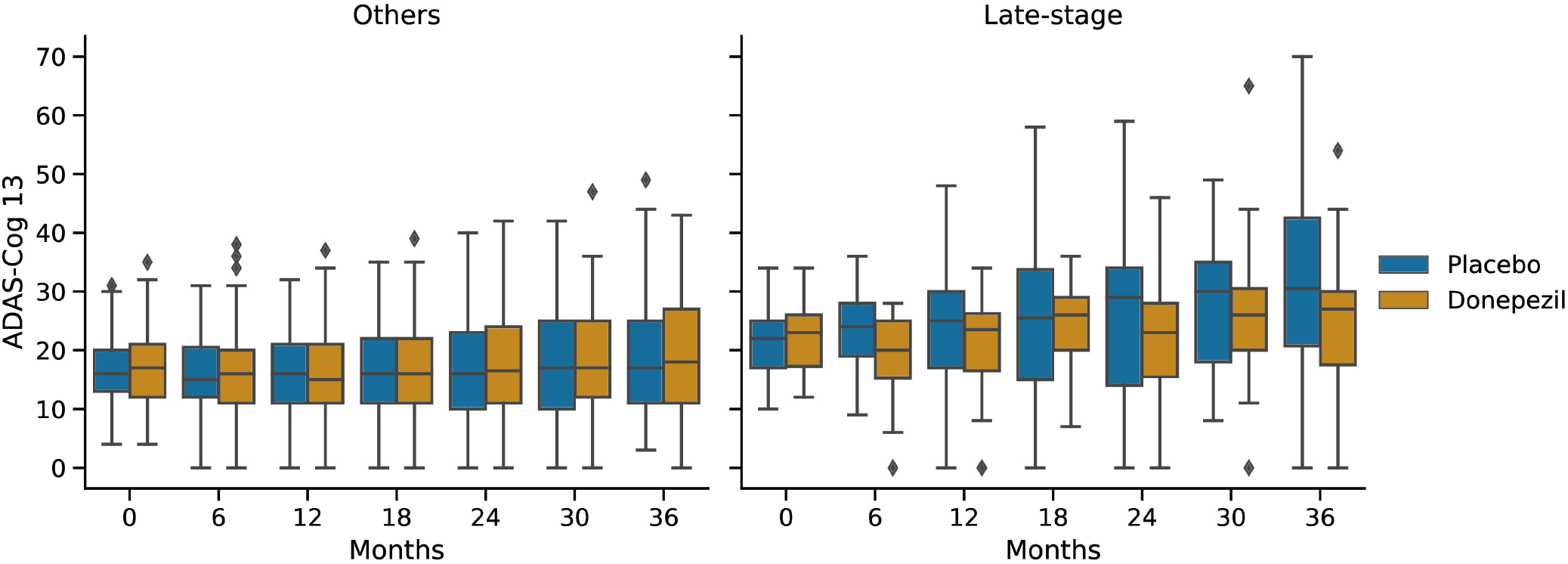
Progression of ADAS-Cog 13 scores in the placebo and donepezil arms throughout the trial for each of the two subgroups.

## 4 DISCUSSION

We fit an event-based model of cognitive decline in Alzheimer’s disease using a reference data set (ADNI), which was then used to score disease progression in subjects at baseline in a completed clinical trial (ADCS-MCI). This disease progression score was used to stratify trial participants for a *post hoc* subgroup analysis of treatment effect.

The event-based model of cognitive decline in Figure 1 is representative of typical (memory-led) Alzheimer’s disease, with CDR and impaired memory recall occurring before decline in verbal fluency, working memory, and executive function. Indeed, the estimated sequence shares similarities with results in (8), which involved an independent cohort. We deliberately excluded ADAS-Cog scores from the model to avoid circularity with the corresponding secondary outcome of the trial (and also to avoid having to perform the relatively arduous ADAS-Cog test at a screening visit). Supplementary Figure 1 shows that the sequence is largely unchanged with ADAS-Cog features included. Notably, Clock Drawing appears as the first event (before even CDR features), albeit with an additional component of positional density around stages 7–9, supporting the presence of additional heterogeneity among individuals. This result warrants further investigation but is beyond the scope of our study.

The event-based model patient staging mechanism (17) revealed considerable heterogeneity in the cognitive impairment of MCI participants in both the ADNI observational study (Figure 3**A**) and the ADCS-MCI clinical trial (Figure 3**B**). Such clinical heterogeneity is likely to mask treatment response in clinical trials, particularly if the underlying source is biological heterogeneity relevant to the experimental treatment. The biological underpinnings here are unknown due to the absence of biomarker data in the ADCS-MCI trial, and we need access to such individual-level biomarker data from more recent clinical trials if we are to assess the value of EBM screening vs biomarker screening. Regardless, we found promising trends in our *post hoc* subgroup analyses (discussed below).

In the ADCS-MCI trial we found encouraging trends towards improved survival (Figure 4), preserved cognition (Figure 6), and a lower hazard ratio (Figure 5) in the more severely affected “Late-stage” MCI subgroup (*N* = 121) compared to the less affected “Others” subgroup (*N* = 648). These results suggest that the treatment (donepezil) may protect cognition and provide more protection against MCI conversion to dementia for late-stage MCI. This result concurs with the fact that donepezil is approved for symptomatic relief in more severely affected groups — specifically, dementia patients. Additionally, a recent re-analysis of the ADCS-MCI trial unmasked beneficial effects of donepezil (26) in a more severely affected subgroup by screening out false-positive MCI participants using hierarchical clustering by Ward’s method.

There are multiple possible explanations for why more severely impaired individuals with MCI seem to benefit from donepezil preferentially over less impaired individuals. For one, donepezil may have less cognitive benefit earlier in the disease. Another is that ADAS-Cog might be inadequate to detect such a benefit. Regardless, the key finding is that our approach was able to stratify a clinical trial population into potential responders and non-responders using only baseline/screening data. This supports the notion that computational, data-driven screening can substantially reduce the size (and cost) of a clinical trial, without sacrificing statistical power (see also Franzmeier et al. (27)).

Our work motivates using event-based model staging as a screening tool to enrich clinical trials, but the general principle can be applied using other models that can calculate disease progression scores, e.g., (28, 29, 30, 31). While many such works mention the potential application to analysing clinical trial data, fewer suggest incorporating this into the screening stage, and none (to our knowledge) have actually applied such models in clinical trials, nor in *post hoc* analyses that follow the original analysis protocol to retrospectively determine subgroup treatment effects. Closest to this work is the aforementioned study of the ADCS-MCI trial data by Edmonds, et al. (26), and the work of Schneider, et al. (32), but the approaches used in these studies do not provide an interpretable disease progression signature, nor do they allow for future extension to seamlessly incorporate imaging data and other biomarkers.

In summary, the ADCS-MCI trial was an attempt to test whether donepezil, an approved symptomatic treatment of dementia patients, could slow progression from MCI to dementia. This placebo-controlled, double-blind, phase 3 study found no significant treatment effects (20). Here we reanalysed the trial in a *post hoc* subgroup analysis, with the subgroups defined by a data-driven disease progression model: the event-based model (8, 17, 19). Our two key findings are: 1) there was considerable heterogeneity in cognitive impairment in the ADCS-MCI trial, suggesting an inadequate screening protocol; 2) this heterogeneity masked a possible treatment effect in a sample of more severely impaired late-stage MCI participants, despite the likelihood of this smaller sample being under-powered to detect an effect of this magnitude. Our study has highlighted a potential mechanism for improving clinical trial design but the general applicability will require broader verification, ideally in more recent trials having biomarker data.

In conclusion, our findings support the use of our proposed data-driven screening method to enhance targeting and efficiency of future clinical trials in Alzheimer’s disease. What is perhaps most exciting in the immediate future is the prospect of performing similar *post hoc* analyses in other “failed” clinical trials, which could resurrect some Alzheimer’s disease drug research programs, saving billions of dollars and years of research. This work is continuing.

## Data Availability

The datasets analysed in this study can be obtained from the respective data controllers: the Alzheimer's Disease Neuroimaging Initiative (ADNI: adni.loni.usc.edu) and Alzheimer's Disease Cooperative Study (ADCS: www.adcs.org). Supporting code for the analyses performed herein can be obtained by contacting the corresponding author.

https://adni.loni.usc.edu

https://www.adcs.org

## AUTHOR CONTRIBUTIONS

NPO conceived of the study and obtained funding. NPO and CS performed the data analysis, and drafted the manuscript. All authors contributed to interpretation of results and manuscript writing.

## ACKNOWLEDGMENTS

The authors are extremely grateful to the participants of the ADNI observational study and ADCS-MCI trial, without whom this research would not have been possible.

Data collection and sharing for this project was funded by the Alzheimer’s Disease Neuroimaging Initiative (ADNI) (National Institutes of Health Grant U01 AG024904) and DOD ADNI (Department of Defense award number W81XWH-12-2-0012). ADNI is funded by the National Institute on Aging, the National Institute of Biomedical Imaging and Bioengineering, and through generous contributions from the following: AbbVie, Alzheimer’s Association; Alzheimer’s Drug Discovery Foundation; Araclon Biotech; BioClinica, Inc.; Biogen; Bristol-Myers Squibb Company; CereSpir, Inc.; Cogstate; Eisai Inc.; Elan Pharmaceuticals, Inc.; Eli Lilly and Company; EuroImmun; F. Hoffmann-La Roche Ltd and its affiliated company Genentech, Inc.; Fujirebio; GE Healthcare; IXICO Ltd.; Janssen Alzheimer Immunotherapy Research & Development, LLC.; Johnson & Johnson Pharmaceutical Research & Development LLC.; Lumosity; Lundbeck; Merck & Co., Inc.; Meso Scale Diagnostics, LLC.; NeuroRx Research; Neurotrack Technologies; Novartis Pharmaceuticals Corporation; Pfizer Inc.; Piramal Imaging; Servier; Takeda Pharmaceutical Company; and Transition Therapeutics. The Canadian Institutes of Health Research is providing funds to support ADNI clinical sites in Canada. Private sector contributions are facilitated by the Foundation for the National Institutes of Health (www.fnih.org). The grantee organization is the Northern California Institute for Research and Education, and the study is coordinated by the Alzheimer’s Therapeutic Research Institute at the University of Southern California. ADNI data are disseminated by the Laboratory for Neuro Imaging at the University of Southern California.

Data collection and sharing for this project was also funded by the Alzheimer’s Disease Cooperative Study (ADCS), funded by the National Institutes of Health Grant U19 AG010483.

The authors acknowledge members of the UCL POND group for valuable discussions of preliminary results.

## SUPPLEMENTAL DATA

Supplementary Material is available in a separate document.

## Supplementary Material

### 1 EVENT-BASED MODEL WITH ADAS-COG

In the main paper we excluded ADAS-Cog from the cognitive instruments used to construct the reference model, allowing separate secondary validation of the subgroups. Here, we provide results with ADAS-Cog included.

#### 1.1 Reference Model

Figure S1 shows the positional variance diagram for the *N* = 803 (of 2040) ADNI participants (229 CN, 177 AD, 397 MCI) with complete data. Compared to Figure 1 in the main paper, the ADAS-Cog features lie grouped together in the middle of the sequence (with the uncertainty around their relative ordering likely driven by their high correlation). The overall sequence is otherwise preserved aside from a swap of the neighbouring DSST and digit span backwards, supporting the robustness of the sequence.

**Figure S1.**
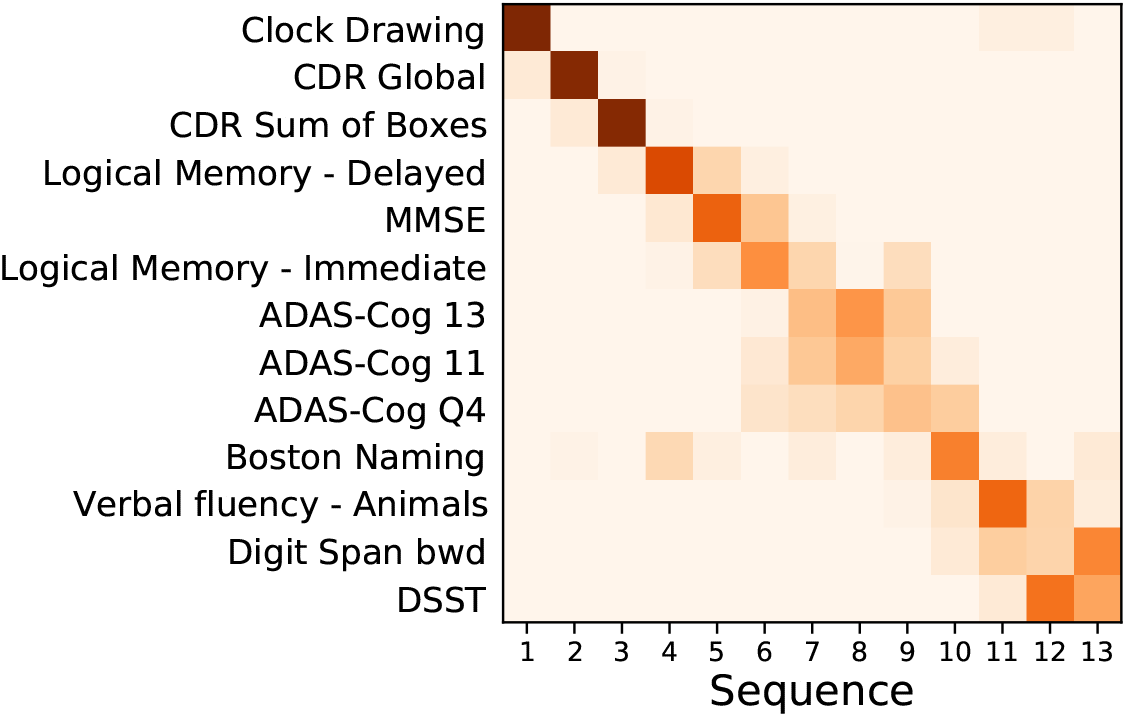
Event based model of cognitive decline (ADNI). Positional density/variance diagram showing the sequence (top to bottom) and uncertainty (left to right) under 5-fold cross-validation (repeated 10 times). Abbreviations: ADAS-Cog — Alzheimer’s Disease Assessment Scale-Cognitive Subscale; CDR — clinical dementia rating; MMSE — mini-mental state examination; bwd — backward; DSST — digit symbol substitution test.

The mixture models corresponding to Figure S1 are shown in Figure S2, which includes ADAS-Cog, highlighting the generally higher scores for AD subjects over those that are cognitively normal. As the mixture models are fit independently for each biomarker, the others are identical to those in the main paper, highlighting the flexibility of this approach to take advantage of additional data where available.

#### 1.2 Patient staging

The model stage distribution for ADCS-MCI trial participants is shown in Figure S3, where (in contrast with the main paper) there are three identifiable subgroups (consisting of 648, 121, and 92 subjects). The late-stage subgroup is still distinct, compromising of the final two stages, though smaller, further corroborating the small difference inclusion of ADAS-Cog had on the disease progression sequence. For completeness, Table S1 provides a group-level comparison between the whole cohort and subgroups, where the most notable difference is in the difference between baseline ADAS-Cog scores in the early- and middle-stage subgroups.

**Figure S2.**
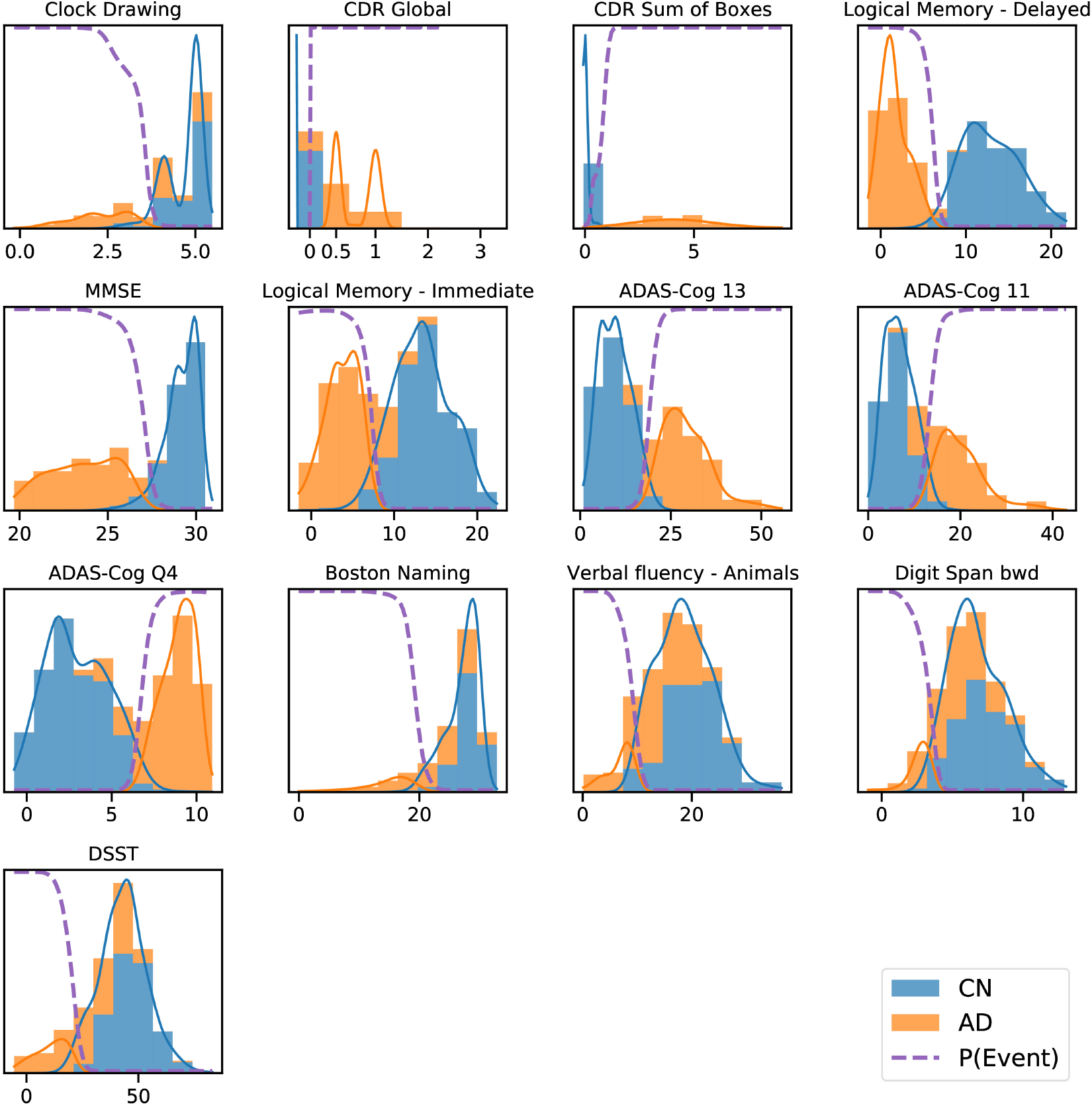
ADNI data histograms (adjusted for age and education level) and EBM mixture models for each feature. Orange bars corresponds to AD patient data, blue bars to data from CN participants, showing the “normal” and “abnormal” distributions and the determined probability of the event having occurred (dashed line).

Kaplan-Meier survival curves are shown in Figure S4 for the whole cohort (**A**), early-stage (**B**), middle-stage (**C**), and late-stage (**D**) subgroups. The late-stage subgroup is similar to that of the main paper (Figure 4**C**), despite the smaller size (92 vs. 121), which is perhaps unsurprising given the similarity between the model sequences with and without ADAS-Cog scores. The lower rate of progression to AD in the early-stage subgroup compared to the middle- and late-stage subgroups supports the validity of the data-driven disease progression sequence, though no subgroup shows a significant difference between placebo and treatment groups.

**Figure S3.**
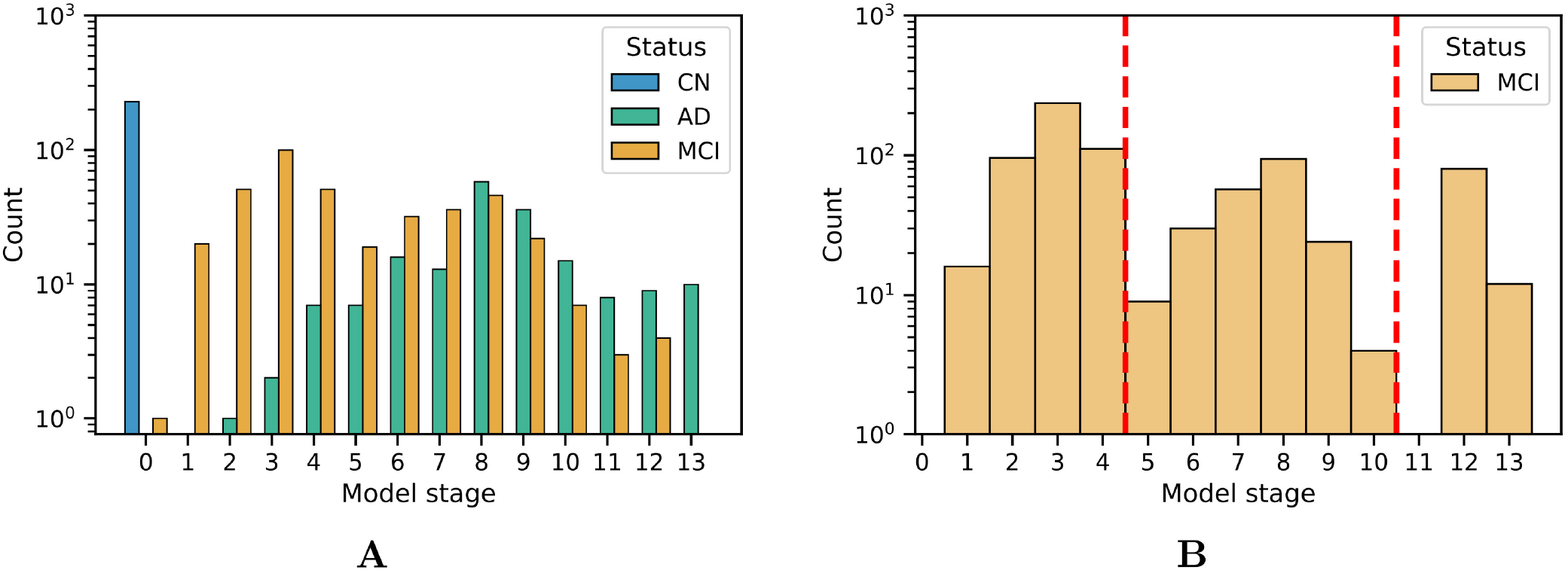
Histograms of model stage for subjects in the ADNI dataset (**A**) and ADCS-MCI trial (**B**).

**Table S1.** Demographic and Cognitive comparison of All ADCS-MCI trial participants and the model-determined subgroups.

| Measure | Group |  |  |  |
| --- | --- | --- | --- | --- |
|  | All<br>(N=769) | Early-stage<br>(N=648) | Middle-stage<br>(N=121) | Late-stage<br>(N=92) |
| Age (years) | 72.9 (7.3) | 72.3 (7.5) | 74.3 (6.5) | 72.7 (7.7) |
| Education (years) | 14.6 (3.1) | 14.9 (3.0) | 13.8 (3.2) | 15.1 (3.1) |
| Sex (% female) | 352 (45.8%) | 197 (42.9%) | 111 (50.9%) | 44 (47.8%) |
| APOE-e4 carrier (%) | 424 (55.1%) | 219 (47.7%) | 146 (67.0%) | 59 (64.1%) |
| Donepezil Arm (%) | 253 (32.9%) | 146 (31.8%) | 81 (37.2%) | 26 (28.3%) |
| Vitamin E Arm (%) | 257 (33.4%) | 152 (33.1%) | 72 (33.0%) | 33 (35.9%) |
| Placebo Arm (%) | 259 (33.7%) | 161 (35.1%) | 65 (29.8%) | 33 (35.9%) |
| ADAS-Cog 11 | 11.3 (4.4) | 8.7 (2.9) | 14.7 (3.6) | 15.6 (3.2) |
| ADAS-Cog 13 | 17.7 (6.1) | 13.8 (3.7) | 23.3 (4.0) | 23.9 (3.9) |
| ADAS-Cog Q4 | 6.3 (2.2) | 5.0 (1.7) | 8.3 (1.3) | 8.0 (1.5) |
| Boston Naming | 6.9 (2.4) | 7.4 (2.2) | 6.7 (2.3) | 5.1 (2.6) |
| CDR Global | 0.5 (0.0) | 0.5 (0.0) | 0.5 (0.0) | 0.5 (0.0) |
| CDR Sum of Boxes | 1.8 (0.8) | 1.6 (0.7) | 2.1 (0.8) | 2.2 (0.8) |
| Clock Drawing | 4.3 (0.9) | 4.5 (0.8) | 4.4 (1.0) | 3.3 (1.0) |
| Digit Span bwd | 6.2 (2.1) | 6.4 (2.1) | 6.2 (2.1) | 5.1 (1.9) |
| DSST | 31.5 (10.9) | 34.7 (10.1) | 29.4 (10.0) | 20.4 (7.9) |
| Logical Memory - Delayed | 3.3 (2.4) | 4.3 (2.2) | 1.8 (1.9) | 1.8 (1.9) |
| Logical Memory - Immediate | 6.2 (3.1) | 7.4 (3.0) | 4.8 (2.6) | 4.3 (2.6) |
| MMSE | 27.3 (1.8) | 28.1 (1.6) | 26.1 (1.4) | 26.0 (1.7) |
| Verbal fluency - Animals | 15.8 (5.2) | 17.4 (5.2) | 14.8 (4.1) | 10.3 (3.1) |

#### 1.3 Summary

ADAS-Cog test scores are commonly-used as a primary endpoint in clinical trials as an assessment of multiple cognitive domains: episodic memory, language, and praxis (Kueper et al., 2018). Excluding it from our reference model (in the main paper) allowed secondary validation of the subgroups, but potentially limited the resolution of the staging that could be obtained. Although cognitive measures (in similar functional domains) will correlate, inclusion of ADAS-Cog into the EBM in these supplementary analyses has allowed further distinction between the early and middle stages to produce three subgroups. The observed hazard ratios indicate similar rates of progression to AD in the donepezil arm, suggesting that (in terms of the primary outcome of the trial) there is not a significant difference in the treatment effect in the early and middle stages of the identified disease progression sequence, despite a consdierable difference in the mean ADAS-Cog 13 at baseline (9.5 points).

**Figure S4.**
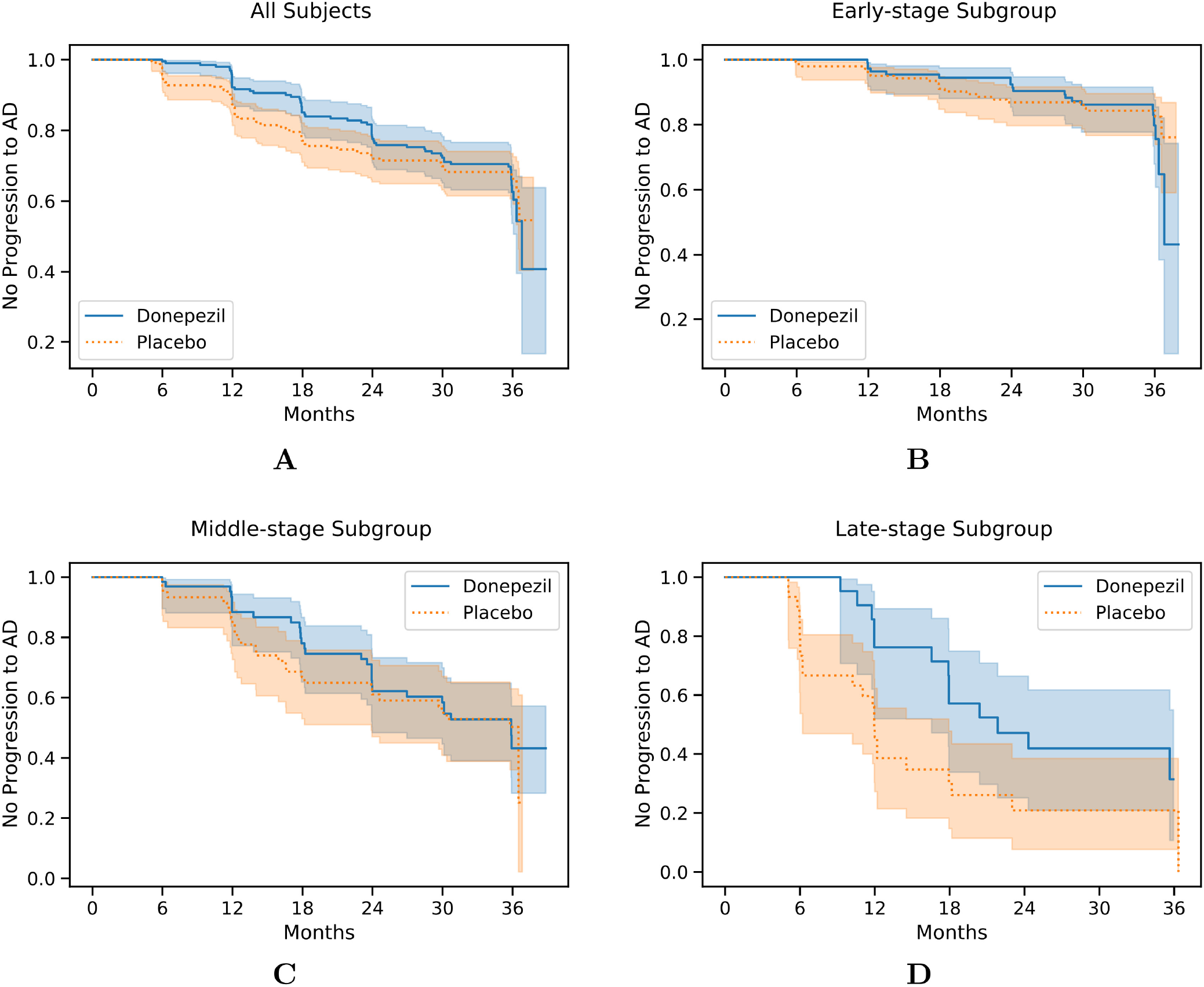
Kaplan-Meier survival curves for all 769 participants (**A**), the early-stage subgroup (**B**), the middle-stage subgroup (**C**), and the late-stage subgroup (**D**) in the ADCS-MCI trial.

Inclusion of ADAS-Cog into the EBM results in a greater separation of mean ADAS-Cog 13 throughout the trial between the treatment arms, though as this is not reflected in the primary outcome this just further supports the current use of donepezil for its cognitive benefits.

## Footnotes

1 Results with ADAS-Cog included can be found in the supplementary material.

## References

1. Cummings J, Ritter A, Zhong K. Clinical Trials for Disease-Modifying Therapies in Alzheimer’s Disease: A Primer, Lessons Learned, and a Blueprint for the Future. Journal of Alzheimer’s Disease 64 (2018) S3–S22. doi:10.3233/JAD-179901. Publisher: IOS Press.

2. Cummings JL, Morstorf T, Zhong K. Alzheimer’s disease drug-development pipeline: few candidates, frequent failures. Alzheimer’s Research & Therapy 6 (2014) 37. doi:10.1186/alzrt269.

3. Cummings J, Morstorf T, Lee G. Alzheimer’s drug-development pipeline: 2016. Alzheimer’s & Dementia: Translational Research & Clinical Interventions 2 (2016) 222–232. doi:10.1016/j.trci.2016.07.001.

4. Cummings J, Lee G, Mortsdorf T, Ritter A, Zhong K. Alzheimer’s disease drug development pipeline: 2017. Alzheimer’s & Dementia: Translational Research & Clinical Interventions 3 (2017) 367–384. doi:10.1016/j.trci.2017.05.002.

5. Cummings J, Lee G, Ritter A, Zhong K. Alzheimer’s disease drug development pipeline: 2018. Alzheimer’s & Dementia: Translational Research & Clinical Interventions 4 (2018) 195–214. doi:10.1016/j.trci.2018.03.009.

6. Cummings J, Lee G, Ritter A, Sabbagh M, Zhong K. Alzheimer’s disease drug development pipeline: 2019. Alzheimer’s & Dementia: Translational Research & Clinical Interventions 5 (2019) 272–293. doi:https://doi.org/10.1016/j.trci.2019.05.008.

7. Cummings J, Lee G, Ritter A, Sabbagh M, Zhong K. Alzheimer’s disease drug development pipeline: 2020. Alzheimer’s & Dementia: Translational Research & Clinical Interventions 6 (2020) e12050. doi:10.1002/trc2.12050.

8. Firth NC, Primativo S, Brotherhood E, Young AL, Yong KX, Crutch SJ, et al. Sequences of cognitive decline in typical alzheimer’s disease and posterior cortical atrophy estimated using a novel event-based model of disease progression. Alzheimer’s & Dementia 16 (2020) 965–973. doi:10.1002/alz.12083.

9. Salloway S, Sperling R, Fox NC, Blennow K, Klunk W, Raskind M, et al. Two phase 3 trials of bapineuzumab in mild-to-moderate alzheimer’s disease. New England Journal of Medicine 370 (2014) 322–333. doi:10.1056/NEJMoa1304839. PMID: 24450891.

10. Wang R, Lagakos SW, Ware JH, Hunter DJ, Drazen JM. Statistics in medicine — reporting of subgroup analyses in clinical trials. New England Journal of Medicine 357 (2007) 2189–2194. doi:10.1056/NEJMsr077003.

11. Cummings J. Lessons learned from alzheimer disease: Clinical trials with negative outcomes. Clinical and Translational Science 11 (2018) 147–152. doi:10.1111/cts.12491.

12. Hardy J, Higgins G. Alzheimer’s disease: the amyloid cascade hypothesis. Science 256 (1992) 184–185. doi:10.1126/science.1566067.

13. Morris GP, Clark IA, Vissel B. Inconsistencies and Controversies Surrounding the Amyloid Hypothesis of Alzheimer’s Disease. Acta Neuropathologica Communications 2 (2014) 135. doi:10.1186/s40478-014-0135-5.

14. Oxtoby NP, Alexander DC, for the EuroPOND consortium. Imaging plus X: multimodal models of neurodegenerative disease. Current Opinion in Neurology 30 (2017). doi:10.1097/WCO.0000000000000460.

15. Marinescu RV, Oxtoby NP, Young AL, Bron EE, Toga AW, Weiner MW, et al. Tadpole Challenge: prediction of longitudinal evolution in Alzheimer’s disease. arXiv (2018).

16. Bilgel M, Jedynak BM, Initiative ADN. Predicting time to dementia using a quantitative template of disease progression. Alzheimer’s & Dementia: Diagnosis, Assessment & Disease Monitoring 11 (2019) 205–215. doi:j.dadm.2019.01.005.

17. Young AL, Oxtoby NP, Daga P, Cash DM, Fox NC, Ourselin S, et al. A data-driven model of biomarker changes in sporadic Alzheimer’s disease. Brain 137 (2014) 2564–2577. doi:10.1093/brain/awu176.

18. Young AL, Marinescu RV, Oxtoby NP, Bocchetta M, Yong K, Firth NC, et al. Uncovering the heterogeneity and temporal complexity of neurodegenerative diseases with Subtype and Stage Inference. Nature Communications 9 (2018) 4273. doi:10.1038/s41467-018-05892-0.

19. Fonteijn HM, Modat M, Clarkson MJ, Barnes J, Lehmann M, Hobbs NZ, et al. An event-based model for disease progression and its application in familial alzheimer’s disease and huntington’s disease. NeuroImage 60 (2012) 1880–1889. doi:10.1016/j.neuroimage.2012.01.062.

20. Petersen RC, Thomas RG, Grundman M, Bennett D, Doody R, Ferris S, et al. Vitamin e and donepezil for the treatment of mild cognitive impairment. New England Journal of Medicine 352 (2005) 2379–2388. doi:10.1056/NEJMoa050151. PMID: 15829527.

21. Jack CR, Knopman DS, Jagust WJ, Shaw LM, Aisen PS, Weiner MW, et al. Hypothetical model of dynamic biomarkers of the alzheimer’s pathological cascade. The Lancet Neurology 9 (2010) 119–128. doi:https://doi.org/10.1016/S1474-4422(09)70299-6.

22. Oxtoby N, Leyland L, Aksman L, Thomas G, Bunting E, Wijeratne P, et al. Sequence of clinical and neurodegeneration events in parkinson’s disease progression. Brain: a journal of Neurology (2021). doi:10.1093/brain/awaa461.

23. Eshaghi A, Marinescu RV, Young AL, Firth NC, Prados F, Jorge Cardoso M, et al. Progression of regional grey matter atrophy in multiple sclerosis. Brain 141 (2018) 1665–1677. doi:10.1093/brain/awy088.

24. Oxtoby NP, Young AL, Cash DM, Benzinger TLS, Fagan AM, Morris JC, et al. Data-driven models of dominantly-inherited Alzheimer’s disease progression. Brain 141 (2018) 1529–1544. doi:10.1093/brain/awy050.

25. Wijeratne PA, Young AL, Oxtoby NP, Marinescu RV, Firth NC, Johnson EB, et al. An image-based model of brain volume biomarker changes in huntington’s disease. Annals of Clinical and Translational Neurology 5 (2018) 570–582. doi:10.1002/acn3.558.

26. Edmonds EC, Ard MC, Edland SD, Galasko DR, Salmon DP, Bondi MW. Unmasking the benefits of donepezil via psychometrically precise identification of mild cognitive impairment: A secondary analysis of the adcs vitamin e and donepezil in mci study. Alzheimer’s & Dementia: Translational Research & Clinical Interventions 4 (2018) 11–18. doi:10.1016/j.trci.2017.11.001.

27. Franzmeier N, Koutsouleris N, Benzinger T, Goate A, Karch CM, Fagan AM, et al. Predicting sporadic alzheimer’s disease progression via inherited alzheimer’s disease-informed machine-learning. Alzheimer’s & Dementia 16 (2020) 501–511. doi:10.1002/alz.12032.

28. Leoutsakos JM, Gross A, Jones R, Albert M, Breitner J. ‘alzheimer’s progression score’: Development of a biomarker summary outcome for ad prevention trials. The Journal of Prevention of Alzheimer’s disease 3 (2016) 229. doi:10.14283/jpad.2016.120.

29. Jedynak BM, Lang A, Liu B, Katz E, Zhang Y, Wyman BT, et al. A computational neurodegenerative disease progression score: Method and results with the alzheimer’s disease neuroimaging initiative cohort. NeuroImage 63 (2012) 1478–1486. doi:https://doi.org/10.1016/j.neuroimage.2012.07.059.

30. Wang Z, Tang Z, Zhu Y, Pettigrew C, Soldan A, Gross A, et al. Ad risk score for the early phases of disease based on unsupervised machine learning. Alzheimer’s & Dementia 16 (2020) 1524–1533. doi:10.1002/alz.12140.

31. Stallard E, Kinosian B, Stern Y. Personalized predictive modeling for patients with Alzheimer’s disease using an extension of Sullivan’s life table model. Alzheimer’s Research & Therapy 9 (2017) 75. doi:10.1186/s13195-017-0302-6.

32. Schneider LS, Frangakis C, Drye LT, Devanand D, Marano CM, Mintzer J, et al. Heterogeneity of treatment response to citalopram for patients with alzheimer’s disease with aggression or agitation: The citad randomized clinical trial. American Journal of Psychiatry 173 (2016) 465–472. doi:10.1176/appi.ajp.2015.15050648.

## References

Kueper JK, Speechley M, Montero-Odasso M. The Alzheimer’s Disease Assessment Scale-Cognitive Subscale (ADAS-Cog): Modifications and Responsiveness in Pre-Dementia Populations. A Narrative Review. J Alzheimers Dis 63 (2018) 423–444.

